# Age at Diagnosis Shapes the Prognosis of Childhood Immune Thrombocytopenia

**DOI:** 10.1101/2020.06.09.20125385

**Authors:** David E. Schmidt, Pernille Wendtland Edslev, Katja M.J. Heitink-Pollé, Rick Kapur, Leendert Porcelijn, C. Ellen van der Schoot, Gestur Vidarsson, Marrie C.A. Bruin, Steen Rosthøj, Masja de Haas

**Affiliations:** Sanquin Research, Department of Experimental Immunohematology, Amsterdam, The Netherlands and Landsteiner Laboratory, Amsterdam UMC, University of Amsterdam, Amsterdam, The Netherlands; Pediatric and Adolescent Health, Division for Oncology and Hematology, Aarhus University Hospital, Aarhus, Denmark; Department of Pediatric Hematology, University Medical Center Utrecht, Utrecht, The Netherlands; Department of Immunohematology Diagnostics, Sanquin Diagnostic Services, Amsterdam, The Netherlands; Princess Maxima Center for Pediatric Oncology, Utrecht, The Netherlands; Department of Pediatrics, Aalborg University Hospital, Aalborg, Denmark; Sanquin Research, Center for Clinical Transfusion Research, Leiden, The Netherlands, and Department of Immunohematology and Blood Transfusion, Leiden University Medical Center, Leiden, The Netherlands

**Keywords:** Immune thrombocytopenia, Paediatrics, Prognosis

## Abstract

**Objective:** Childhood immune thrombocytopenia (ITP), an acquired bleeding disorder, occurs at any age. Studies have indicated a less favourable prognosis in children aged above ten years. Low lymphocyte counts have been proposed as predictors of chronic disease. Detailed knowledge of ITP disease characteristics and prognosis at various ages may be useful to support clinical decision-making. We aimed to define how age shapes the clinical characteristics, biological parameters and disease outcomes in childhood ITP.

**Design:** Post-hoc analysis of two prospective European studies (NOPHO ITP study and TIKI trial). Children were followed for 6-12 months.

**Setting:** Patient inclusion in paediatrics departments in the Netherlands and the five Nordic countries.

**Patients:** Children aged <16 years with newly diagnosed ITP (N=577) and severe thrombocytopenia (diagnosis platelet count ≤20 × 10^9^/L).

**Results:** By analysing age effects on a continuous scale, we observed that recovery rates at 3-12 months follow-up were gradually reduced in children aged above five years. An absence of a response to IVIg was observed at all ages, but was more common in older children, in particular above 6 years of age. Leukocyte and lymphocyte subset counts were reduced with age, but not elevated or decreased compared to age-appropriate reference intervals. Children aged below seven years showed elevated thrombopoietin levels.

**Conclusions:** Already from five years of age onwards, there is an increasing risk for a long-lasting course of ITP. Given the varying treatment responses and biological variation, age differences should be considered for the design of clinical trials, prediction models and biological studies.

## INTRODUCTION

Childhood ITP is a rare acquired bleeding disorder, characterized by isolated thrombocytopenia (platelet count <100 × 10^9^/L) that is not explained by alternative causes. [1] The morbidity of ITP is most pronounced in children with persistent and chronic disease courses.[2,3] In 80-90% of children, ITP resolves within one year after diagnosis.[4-6] Although ITP shows a predisposition to young children with a median age of 4 years,[4,5,7] children and adolescents at all ages can be affected.[7,8] Paediatricians are experts in taking the patient’s age into account in their clinical reasoning (i.e., for diagnosis, treatment, and prognosis). This requires detailed knowledge of age-dependent disease characteristics.

To date, the clinical presentation and the prognosis of ITP at various ages are incompletely defined. Studies on the identification of predictors of ITP disease courses have not always been adjusted for age or used an arbitrary definition and categorization of age groups, often in dichotomous strata.[8-12] In a systematic review, combined effect estimates of 13 studies supported that children ≥11 years at diagnosis are of higher risk of chronic ITP, and four studies further suggested that such effects are already present with an age ≥8 years.[12] The most comprehensive available data from the ICIS study showed that older children above 10 year also exhibited a reduced recovery chance during follow-up, compared to younger children.[8,10] These findings have been corroborated by independent studies.[11,13] Concerning treatment outcomes, a cohort study in 25 children suggested that a platelet response to corticosteroid and IVIg might be less likely in children above the age of 10 years.[14] Altogether, there are data supporting divergent clinical characteristics and disease outcomes at various ages, but a detailed evaluation of these age-dependent differences with regard to the clinical presentation and the prognosis of ITP is lacking.

In the present study, we aimed to describe in detail the age-dependent changes in the clinical presentation, biological profile and disease outcomes of newly diagnosed ITP. Two large European multicentre studies that both assessed longitudinal disease outcomes for 6-12 months were combined (N=577). In addition to the long-term follow-up, extensive biological data were available, and the effects of age on IVIg treatment responses could be assessed.

## METHODS

### Study subjects and ethics statement

Data from two prospective multicentre studies were available for this study. Briefly, in the Nordic Pediatric Hematology-Oncology (NOPHO) ITP study, 377 children with newly diagnosed ITP with an age <15 years and a presenting platelet count ≤20 × 10^9^ / L were included in a prospective registry study in the five Nordic countries and followed for six months for disease outcomes.[15] Study approval was obtained from the ethical review committees and authorities in each participating country. In the *Therapy With or Without IVIg in Kids with ITP* (TIKI) trial,[4] 200 children with newly diagnosed ITP with an age between 3 months and ≤16 years and a presenting platelet count ≤20 × 10^9^/L were included in a randomized controlled trial in 46 hospitals in The Netherlands. Children with prior immunotherapy (within three months) or severe or life-threatening bleeding were excluded. The children were followed for one year for disease outcomes. Data was collected on standardized clinical report forms. The study was approved by the ethical review committee of University Medical Center Utrecht, appropriate informed consent was obtained as described.[4] All analyses were performed on unidentified, coded data.

### Clinical outcome definition

Bleeding symptoms were scored according to the modified Buchanan scale.[16,17] Recovery was evaluated according to outcome criteria by the International Working Group;[1] a complete recovery was defined as a platelet count ≥100 × 10^9^ /L (TIKI) and ≥150 × 10^9^ /L (NOPHO). Transient ITP was defined as a complete recovery three months after diagnosis;[18] chronic ITP was defined as absence of a complete recovery twelve months after diagnosis.[1] Response to IVIg was evaluated by the same recovery criteria. A sustained response to IVIg was defined as a response with a platelet count ≥100 × 10^9^ /L, that was present one week after treatment and was sustained at the next evaluation, i.e. one month after treatment, as described by the IWG.[1]

### Laboratory analyses

Additional laboratory analyses were available for the TIKI trial. Complete blood count data were collected from routine clinical chemistry at the study centre through clinical report forms. The presence of circulating anti-platelet antibodies was evaluated by monoclonal antibody-specific immobilization of platelet antigens (MAIPA; glycoprotein-specific) as previously described.[19] Lymphocyte subset immune phenotyping was performed in a centralized laboratory by flow cytometry (Sanquin Immunodiagnostics), using multicolour staining (BD Biosciences; Vianen, The Netherlands; #644611) in TruCount tubes (BD Biosciences #340334) on heparinized whole blood. Thrombopoietin was measured using an in-house ELISA[20,21] and expressed in arbitrary units (AU/mL), where one AU is equal to 9 pg of recombinant TPO (Research Diagnostics, Flanders, NJ, USA).

### Statistical analysis

Data were analysed in R version 3.6.1 (R Core Team). The effect of age was determined using linear and logistic regression, for continuous and categorical variables, respectively.

### Data sharing

Data may be requested from the corresponding author for academic collaborations.

## RESULTS

A total of 577 children with newly diagnosed ITP were available for analysis, 377 from the NOPHO ITP study and 200 from the TIKI trial (Table 1). The age distributions in both cohorts were similar (Figure 1A), with the 25^th^, 50^th^ and 75^th^ percentile at two, four and seven years of age, respectively. Both cohorts showed a comparable proportion of girls, children with insidious disease onset, preceding infection or vaccination, as well as a similar frequency of recovery during follow-up (Table 1).

**Table 1.** Baseline characteristics of the study cohorts at the time of diagnosis.

|  | **TIKI (N=200)** | **NOPHO (N=377)** |
| --- | --- | --- |
| Age, years | 4.0 [2.4 – 7.5] | 4.0 [2.0 – 7.0] |
| Female | 91 (46%) | 162 (43%) |
| Mucosal bleeding | 80 (40%) | 165 (44%) |
| Insidious disease onset | 30 (15%) | 84 (22%) |
| Preceding infection | 108 (55%) | 217 (58%) |
| Preceding vaccination | 8 (4%) | 27 (7%) |
| Platelet count, $\times 10^9/\text{L}$ | 6 [3 – 10] | 7 [4 – 12] |
| Leukocytes, $\times 10^9/\text{L}$ | 8.3 [6.6 – 10.6] | |
| Recovery at 3 months | 144 (73%) | 255 (67%) |
| Recovery at 6 months | 162 (81%) | 282 (75%) |
| Recovery at 12 months | 179 (89%) |  |

Data are median [IQR] or count (%).
Insidious disease onset, symptoms $\geq 14$ days at the time of diagnosis.
Preceding infection and vaccination within the prior 28 days.

**Figure 1.**
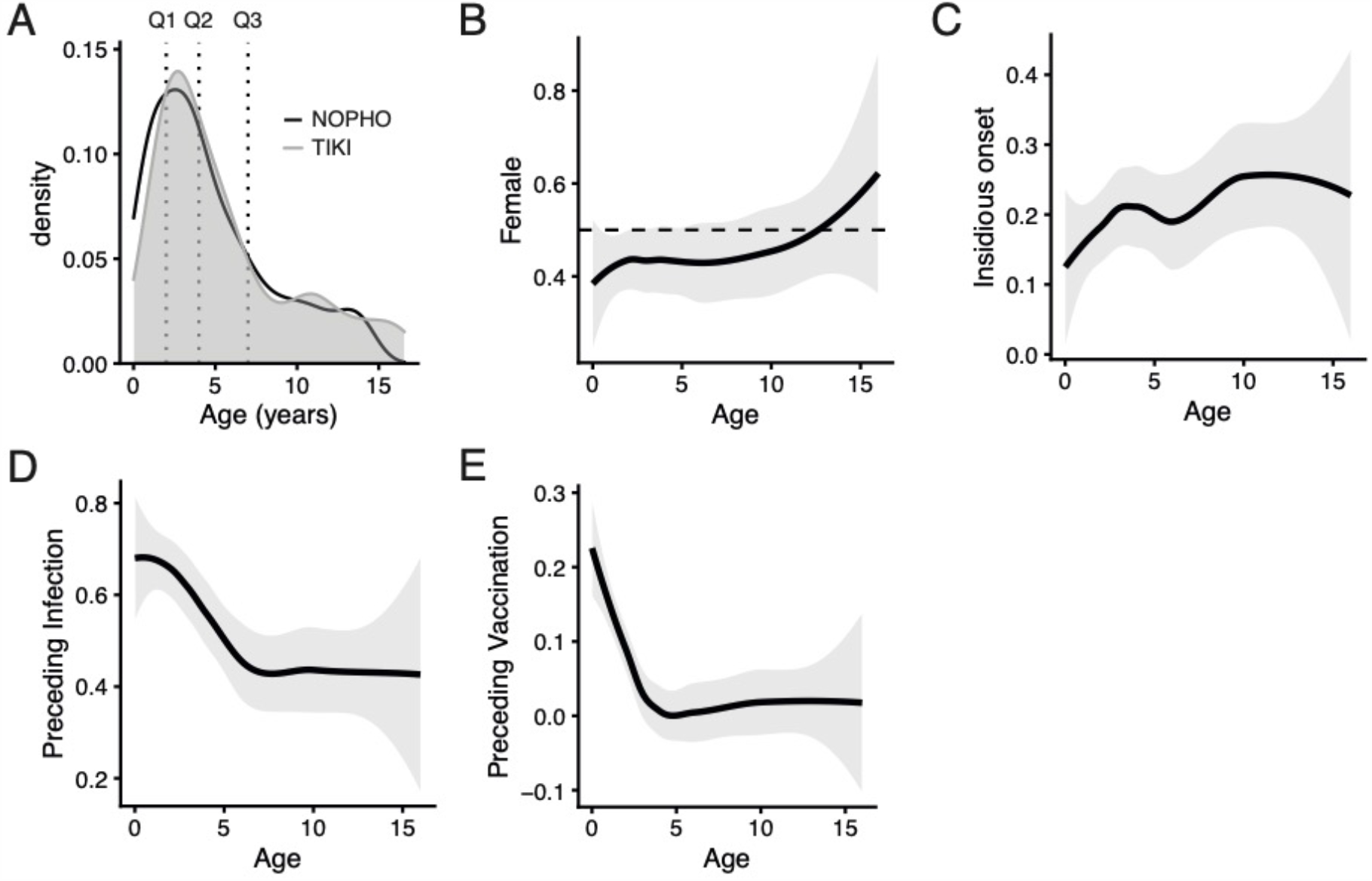
Clinical characteristics by age at presentation. (A) Both cohorts, TIKI (N=200; grey area) and NOPHO (N=377; black line), show identical age distribution. The 25^th^, 50^th^, and 75^th^ quantiles (Q1-Q3) were 2, 4 and 7 years. (B-E) Proportion of females, insidious disease onset (symptoms > 14 days at the time of diagnosis), preceding infection or vaccination (within 28 days). Depicted are locally estimated scatterplot smoothing (LOESS) regression curves (black line) with corresponding standard errors (grey area).

### Clinical presentation correlates with age at diagnosis

Boys were slightly overrepresented amongst children with newly diagnosed ITP (Table 1). With higher age at presentation, the proportion of girls increased (Figure 1B); girls were overrepresented in children above 12 years of age at presentation. An insidious disease onset, defined as symptoms >14 days, was uncommon in the youngest children, and became more prevalent with increasing age at diagnosis (Figure 1C). Mucosal bleeding was present with the same proportion at all ages (data not shown). Many cases of childhood ITP occur after a preceding infection or vaccination. We observed that almost 70% of young children had a self-reported history of a preceding infection; this dropped to about 40% in children in children aged above five years (Figure 1D). In line with the routine national vaccination schemes, a preceding vaccination was present in children below four years of age (Figure 1E).

### Recovery from ITP

The probability of recovery three months after diagnosis (i.e., transient ITP) was consistently above 70% in children below five years of age, and gradually decreased to below 60% with increasing age at presentation (Figure 2A, upper panel). The odds ratio for recovery from ITP given a one-year increase in age was 0.93 (95% CI, 0.89 – 0.97). Notably, in absolute numbers, most of the children without recovery were below five years of age (Figure 2A, lower panel). A similar decrease in the proportion of recovery six months after the diagnosis was observed with increasing age at diagnosis (Figure 2B, upper panel). The odds ratio for recovery from ITP at six months, with a one-year increase in age, was 0.92 (95% CI, 0.88 – 0.97). The probability of chronic ITP, i.e., absence of recovery twelve months after the diagnosis, in children younger than five years was below 5%, and increased to 20% in older children (Figure 2C, upper panel). Taken together, these data showed that children as young as five years of age at the diagnosis had already a less favourable prognosis than younger children.

**Figure 2.**
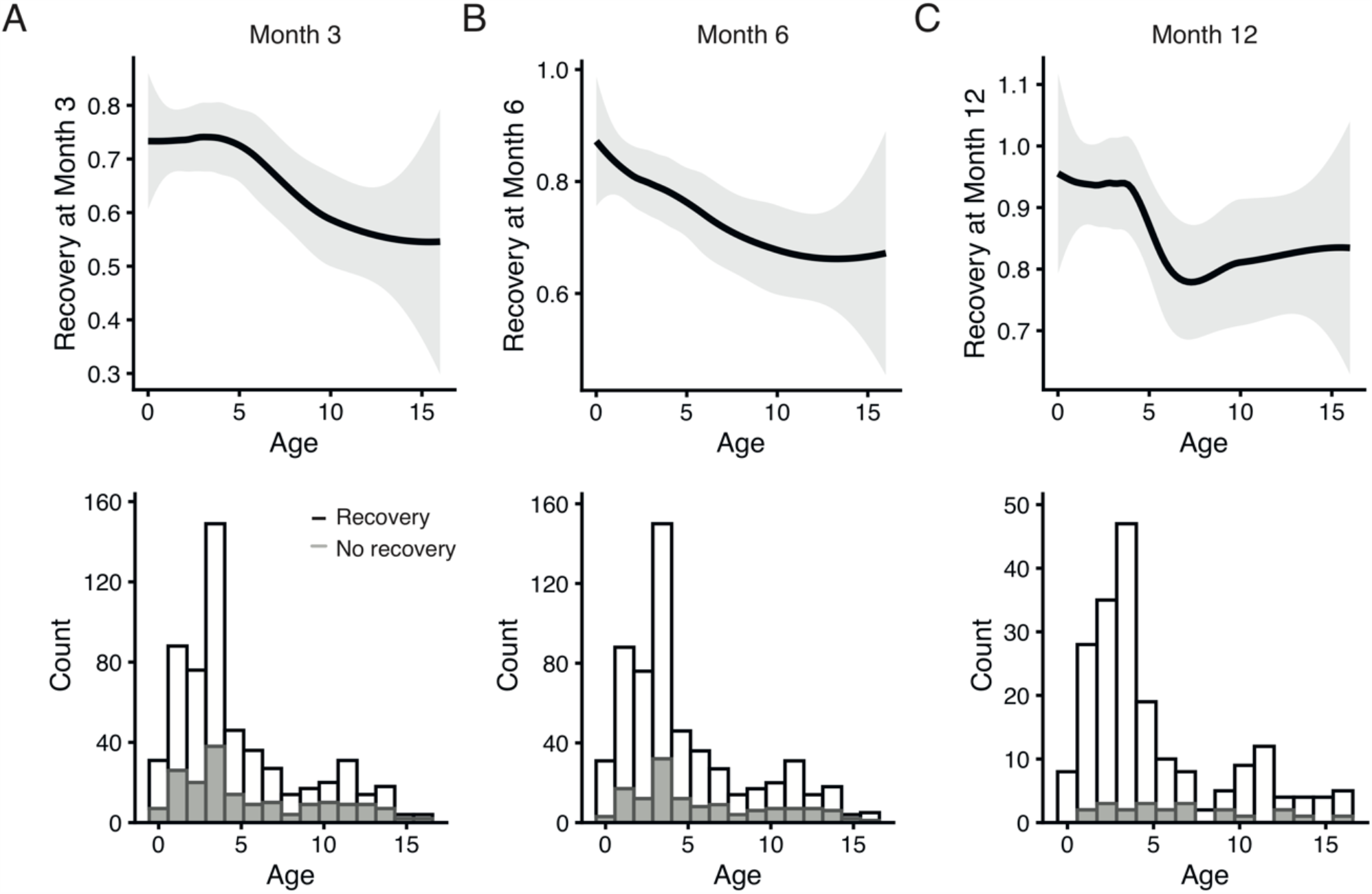
Relationship between ITP disease outcomes and age at presentation. Upper panel, recovery from ITP per age at presentation. Depicted are locally estimated scatterplot smoothing (LOESS) regression curves (black line) with corresponding standard errors (grey area). Lower panel, histogram for children who recovered *versus* did not recover at the timepoints. Data for month 3 and month 6 after diagnosis were available for NOPHO and TIKI studies, data for month 12 was only available from the TIKI trial.

### Response to IVIg treatment

For children in the TIKI trial that were randomized to receive IVIg treatment (N=100), we assessed complete sustained responses (CSR) to IVIg, i.e., a platelet response ≥100 × 10^9^/L one week after IVIg that is sustained one month after the treatment. Overall, CSR was present in 0.57 (n/N, 56/99; 95% CI, 0.47 – 0.66); the complete response frequency one week after IVIg treatment was 0.69 (n/N, 68/99; 95% CI, 0.60 - 0.78). Importantly, we observed that non-responders to IVIg were present among all age groups (Figure 3). However, a complete sustained response after IVIg treatment was less probable with increasing age, showing a bimodal distribution (Figure 3). On average, a one-year increase in age had an odds ratio for a CSR to IVIg of 0.90 (95% CI, 0.80 – 1.00). Being aware that there was limited data available for children above 10 years, the response rate of patients aged 10 years or older was only 0.29 (n/N, 4/14; 95% CI, 0.05 – 0.52); none of the six patients above the age of 12 showed a complete sustained response. In sum, whereas children at all ages showed absence of a sustained recovery after IVIg, the chance to recover after treatment was reduced with increasing age at diagnosis.

**Figure 3.**
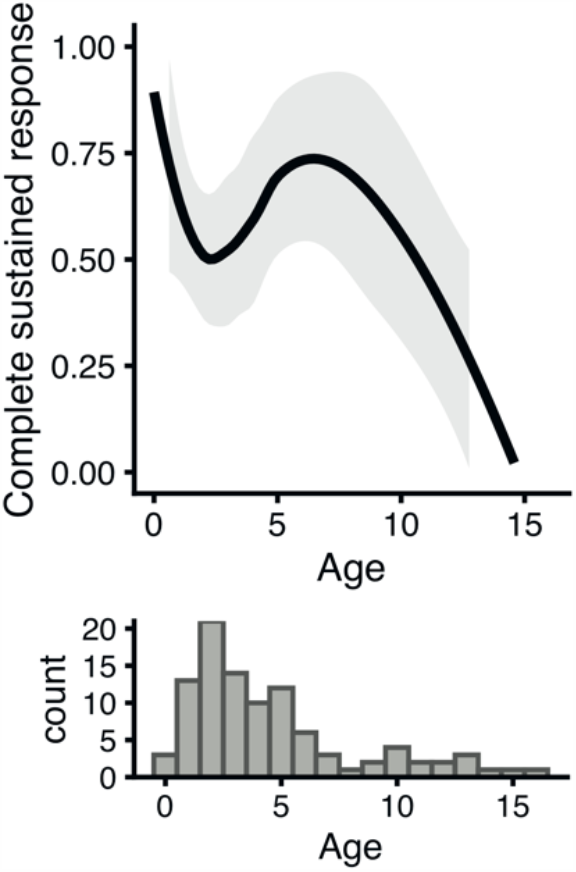
Response to IVIg stratified by age for children in the treatment arm of the TIKI trial (N=100). Depicted is a locally estimated scatterplot smoothing (LOESS) regression curves (black line) with corresponding standard errors (grey area). The lower panel shows a histogram of the available observations per age; in particular for children ≥7 years of age there were few observations and estimates should be interpreted with caution.

### The biological profile correlates with age at presentation

Finally, we assessed the relationship of age at diagnosis with laboratory parameters that have been implicated in the ITP pathophysiology,[22] or proposed as candidate prediction markers for disease outcomes.[12] Of note, a platelet count of ≤20 × 10^9^/L was required for study inclusion. The youngest children showed higher platelet counts at diagnosis, but overall the platelet count was unaltered with age (Figure 4A). The absolute leukocyte and lymphocyte counts decreased with increasing age at presentation until ∼6-8 years of age, and then showed a consistent mean (Figure 4B-C). These changes were fully in line with the age-matched reference ranges (dotted lines, Figure 4).[23,24] We observed the same when the respective T and B cell subsets of CD4, CD8, and CD19 cells were assessed (Figure 4D-F). NK cell counts were moderately higher in children with an age below 5 years at diagnosis (Figure 4G), compared to older children, in agreement with appropriate reference range. Thrombopoietin (TPO) decreased with age until ∼5 years of age (Figure 4H), which is the same trend of age-associated changes shown in published reference data.[25] However, although TPO showed the same age-related changes as observed in healthy individuals, the TPO levels were increased relative to the reference intervals of healthy controls (adult 97.5^th^ percentile, 32 AU/mL) [20], in particular in the younger children (Figure 4H). Haemoglobin levels were increased with age, within the age-appropriate reference ranges (Figure 4I). Contrary to a potential correlation with higher preceding infection rates, C-reactive protein levels were not different at various ages (N=79 consecutive samples; data not shown).

**Figure 4.**
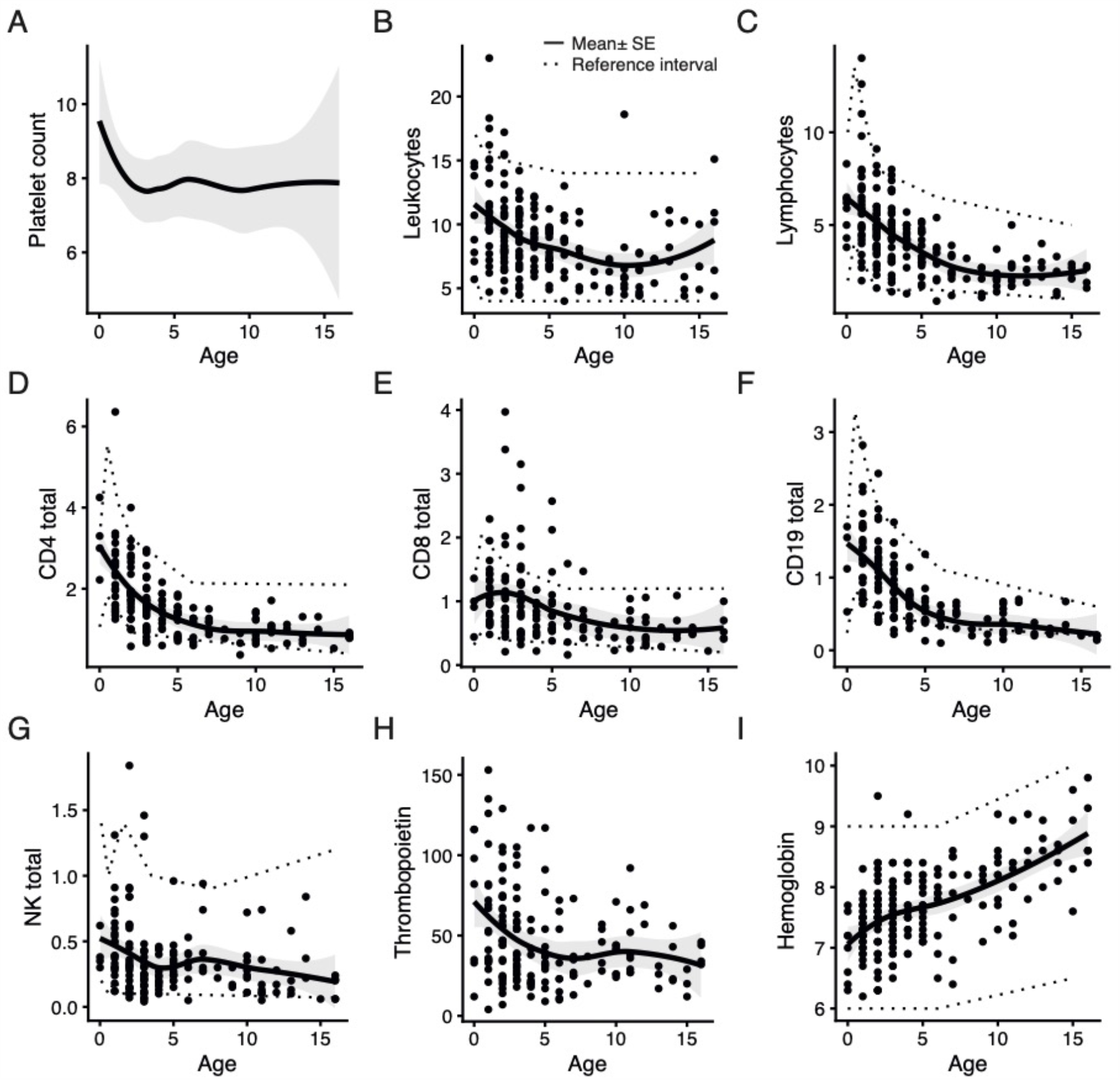
Cell frequencies and plasma markers by age at presentation. (A-C) Cell counts by routine analysis; all data are × 10^9^/L. In (A), data are shown for TIKI and NOPHO cohorts (N=577); for (B-C) data are only shown for the TIKI cohort (N=200). Dotted lines indicate the 2.5^th^ and 97.5th percentiles. (D-G) Absolute cell counts by immune phenotyping in the TIKI study (N=159). T cells (CD4, CD8), B cells (CD19) and NK cells (CD3- CxD56+ CD16+) were determined using flow cytometry. All data are × 10^9^/L. Dotted lines indicate the 5^th^ and 95th percentiles.[23,24] (H) Plasma thrombopoietin levels (AU/mL) as determined by ELISA (N=168). The 97.5^th^ reference percentile is 32 AU/mL. Not shown is one individual with a TPO of 693 AU/mL and age at diagnosis of 5 years. (I) Haemoglobin levels (mmol/L) per age (N=197). Depicted are locally estimated scatterplot smoothing (LOESS) regression curves (black line) with corresponding standard errors (grey area).

The frequency of circulating IgM anti-platelet glycoprotein IIb/IIIa, Ib/IX and V antibodies decreased with age (Figure 5A), with an odds ratio of 0.85 (95% CI, 0.79 – 0.92) for a one-year increase in age. Moreover, although there seemed to be a lower frequency of IgG anti-platelet glycoprotein antibodies in the oldest children (Figure 5B), our data were inconclusive about this effect with an odds ratio of 0.93 (95% CI, 0.80 – 1.06) for a one-year increase in age.

**Figure 5.**
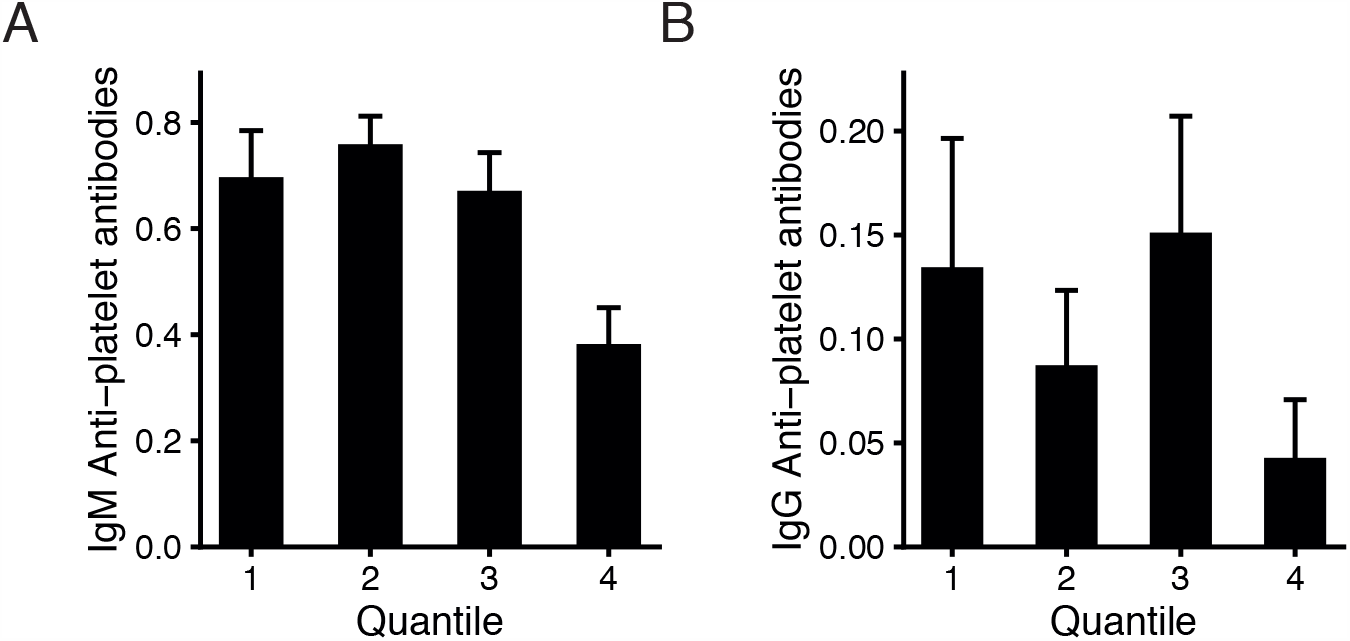
Anti-platelet antibodies in the TIKI cohort for the various age quantiles, for (A) IgM (N=167) and (B) IgG (N=176). Antibodies were determined by glycoprotein-specific analysis for GP Ib/IX, IIb/IIIa and V using MAIPA, as previously described.[19] The 25^th^, 50^th^, and 75^th^ quantiles (Q1-Q3) were 2, 4 and 7 years.

## DISCUSSION

The main findings of our study are that below five years of age, the clinical presentation and disease outcomes are favourable and relatively homogeneous. This is a younger age than previously acknowledged with the arbitrarily chosen eight- or ten-years thresholds.[8,12] Thereafter, the prognosis is increasingly unfavourable with advancing age, and may represent a different disease spectrum. Despite the favourable recovery in children aged below five years, this age group still accounts for most cases of persistent ITP and of non-responders to IVIg.

It is of key importance that the association of the ITP disease characteristics and prognosis with age is considered during the clinical assessment and for the design of clinical and molecular prediction models. The lack of a response to IVIg with increasing age suggests that other pathophysiological mechanisms could be relevant for this older age group. It also implies that clinical trials aiming to evaluate novel treatments for ITP should be designed to stratify children by age, to not miss potential benefits.

Our study also highlights the importance of adjustment of candidate prediction markers of disease outcomes for age, due to potential confounding. When such markers are considered as predictors of e.g. transient vs. chronic ITP, without adjustment for age, it may falsely be assumed that they are associated with disease outcomes. In this study, the major lymphocyte subsets all changed with age. Although the assessed lymphocyte counts were all within the appropriate reference ranges for the respective age groups.

### Findings of others

Childhood ITP may be different from adult ITP in terms of clinical characteristics, biological parameters and disease outcomes,[26,27] but at what age do such differences start to arise? Clearly, infants (up to 1 year of age) have been previously reported to have favourable disease outcomes,[8,10] and other studies showed that this favourable relationship was also present in children up to two or three years of age.[28,29] On the other side of the age spectrum, Lowe and Buchanan described that adolescents with childhood ITP share features of childhood and adult ITP,[9] using an arbitrary threshold for analysis of ten years of age. In a systematic review, pooled effects of several studies showed that an age at diagnosis ≥11 years was a predictor of chronic ITP,[12] and a small set of studies suggested that this effect may be present already as of 8 years of age. We extend the findings of these previous studies by evaluating a large and unselected population, and analysing age effects on a continuous scale, thus showing that differences in the clinical presentation and prognosis diverge around five years of age.

### Strengths and Limitations

A key strength of this study is the data from two multicentre and multinational studies. These studies selected children in paediatric departments at the time of diagnosis, without demonstrable follow-up bias.[5] As such, the study population is reflective of the unselected childhood ITP cases, without bias to older children or more severe presentations that may be seen at academic centres. Moreover, we had a relatively large study population of 577 children available for analysis, and both cohorts exhibit similar baseline characteristics and longitudinal follow-up. Follow-up data beyond six months were only available for the TIKI cohort. Our study’s generalizability is limited by the inclusion of patients with a platelet count ≤20 × 10^9^/L, reflecting the most severe ITP presentations. These children were chosen in the studies to define a population at risk of bleeding and potential need of treatment.

## Conclusions

In conclusion, our study defines a threshold with changes in the clinical presentation and disease outcomes with childhood ITP from five years of age. Lymphocyte subsets are affected by age, and follow the age-appropriate reference ranges. For the clinical management of childhood ITP, such as the consideration of treatment or additional diagnostic investigations, age at diagnosis should be taken into account. Importantly, age is likely an important confounder in biological studies and should be considered during the design of clinical trials and prediction models.

## Data Availability

Data may be requested from the corresponding author for academic collaborations.

## Authorship Contributions

DES designed the study, analysed and interpreted data, and wrote the manuscript. KMJHP, PWE, MCAB and SR contributed clinical data, supervised analyses, interpreted data, and discussed results. RK, GV, CEvdS and LP interpreted data and discussed results. MdH designed and supervised the study. All co-authors reviewed, revised and approved the manuscript.

## References

1. Rodeghiero F, Stasi R, Gernsheimer T, Michel M, Provan D, Arnold DM, et al. Standardization of terminology, definitions and outcome criteria in immune thrombocytopenic purpura of adults and children: report from an international working group. American Society of Hematology; 2009 Mar 12;113(11):2386–93.

2. Heitink-Polle KMJ, Haverman L, Annink KV, Schep SJ, de Haas M, Bruin MCA. Health-related quality of life in children with newly diagnosed immune thrombocytopenia. Haematologica. 2014 Aug 31;99(9):1525–31.

3. Flores A, Klaassen RJ, Buchanan GR, Neunert CE. Patterns and influences in health-related quality of life in children with immune thrombocytopenia: A study from the Dallas ITP Cohort. Pediatr Blood Cancer. John Wiley & Sons, Ltd; 2017 Aug;64(8):e26405.

4. Heitink-Pollé KMJ, Uiterwaal CSPM, Porcelijn L, Tamminga RYJ, Smiers FJ, van Woerden NL, et al. Intravenous immunoglobulin vs observation in childhood immune thrombocytopenia: a randomized controlled trial. 2018 Aug 30;132(9):883–91.

5. Rosthøj S, Hedlund-Treutiger I, Rajantie J, Zeller B, Jonsson OG, Elinder G, et al. Duration and morbidity of newly diagnosed idiopathic thrombocytopenic purpura in children: A prospective Nordic study of an unselected cohort. J Pediatr. 2003 Sep;143(3):302–7.

6. Bennett CM, Neunert C, Grace RF, Buchanan G, Imbach P, Vesely SK, et al. Predictors of remission in children with newly diagnosed immune thrombocytopenia: Data from the Intercontinental Cooperative ITP Study Group Registry II participants. Pediatr Blood Cancer. 2017 Aug 9;65(1):e26736–7.

7. Kuhne T, Imbach P, Bolton-Maggs PH, Berchtold W, Blanchette V, Buchanan GR, et al. Newly diagnosed idiopathic thrombocytopenic purpura in childhood: an observational study. Lancet. 2001 Dec;358(9299):2122–5.

8. Kühne T, Buchanan GR, Zimmerman S, Michaels LA, Kohan R, Berchtold W, et al. A prospective comparative study of 2540 infants and children with newly diagnosed idiopathic thrombocytopenic purpura (ITP) from the Intercontinental Childhood ITP Study Group. J Pediatr. 2003 Nov;143(5):605–8.

9. Lowe EJ, Buchanan GR. Idiopathic thrombocytopenic purpura diagnosed during the second decade of life. J Pediatr. 2002 Aug;141(2):253–8.

10. Imbach P, Kuhne T, Müller D, Berchtold W, Zimmerman S, Elalfy M, et al. Childhood ITP: 12 months follow-up data from the prospective registry I of the Intercontinental Childhood ITP Study Group (ICIS). Pediatr Blood Cancer. 2006 Mar;46(3):351–6.

11. Donato H, Picón A, Martinez M, Rapetti MC, Rosso A, Gomez S, et al. Demographic data, natural history, and prognostic factors of idiopathic thrombocytopenic purpura in children: a multicentered study from Argentina. Pediatr Blood Cancer. Wiley Subscription Services, Inc., A Wiley Company; 2009 Apr;52(4):491–6.

12. Heitink-Polle KMJ, Nijsten J, Boonacker CWB, de Haas M, Bruin MCA. Clinical and laboratory predictors of chronic immune thrombocytopenia in children: a systematic review and meta-analysis. 2014 Nov 20;124(22):3295–307.

13. Rosthøj S, Hedlund-Treutiger I, Rajantie J, Zeller B, Jonsson OG, Henter JI. Age-dependent differences in Nordic children with ITP. Journal of Pediatrics. 2005 Jan;146(1):151–2.

14. Ho W-L, Lee C-C, Chen C-J, Lu M-Y, Hu F-C, Jou S-T, et al. Clinical features, prognostic factors, and their relationship with antiplatelet antibodies in children with immune thrombocytopenia. J Pediatr Hematol Oncol. 2012 Jan;34(1):6–12.

15. Edslev PW, Rosthøj S, Treutiger I, Rajantie J, Zeller B, Jonsson OG, et al. A clinical score predicting a brief and uneventful course of newly diagnosed idiopathic thrombocytopenic purpura in children. British Journal of Haematology. Blackwell Publishing Ltd; 2007 Aug;138(4):513–6.

16. Bennett CM, Rogers ZR, Kinnamon DD, Bussel JB, Mahoney DH, Abshire TC, et al. Prospective phase 1/2 study of rituximab in childhood and adolescent chronic immune thrombocytopenic purpura. 2006 Apr 1;107(7):2639–42.

17. Buchanan GR, Adix L. Grading of hemorrhage in children with idiopathic thrombocytopenic purpura. J Pediatr. 2002 Nov;141(5):683–8.

18. Schmidt DE, Heitink-Pollé KMJ, Laarhoven AG, Bruin MCA, Veldhuisen B, Nagelkerke SQ, et al. Transient and chronic childhood immune thrombocytopenia are distinctly affected by Fc-γ receptor polymorphisms. Blood Adv. 2019 Jul 9;3(13):2003–12.

19. Schmidt DE, Heitink-Pollé KMJ, Porcelijn L, van der Schoot CE, Vidarsson G, Bruin MCA, et al. Anti-Platelet Antibodies in Childhood Immune Thrombocytopenia: Prevalence and Prognostic Implications. J Thromb Haemost. 2020 Feb 13;:jth.14762.

20. Folman CC, Borne von dem AE, Rensink IH, Gerritsen W, van der Schoot CE, de Haas M, et al. Sensitive measurement of thrombopoietin by a monoclonal antibody based sandwich enzyme-linked immunosorbent assay. Thromb Haemost. 1997 Oct;78(4):1262–7.

21. Porcelijn L, Schmidt DE, van der Schoot CE, Vidarsson G, de Haas M, Kapur R. Antiglycoprotein Ibα autoantibodies do not impair circulating thrombopoietin levels in immune thrombocytopenia patients. Haematologica. 2019 Jul 11;:haematol.2019.228908.

22. Zufferey A, Kapur R, Semple JW. Pathogenesis and Therapeutic Mechanisms in Immune Thrombocytopenia (ITP). J Clin Med. 2017 Feb 9;6(2):16.

23. van den Heuvel PhD D, MD MAEJ, MSc KN, PhD WAD, van Lochem PhD EG, PhD LEB-J, et al. Effects of nongenetic factors on immune cell dynamics in early childhood: The Generation R Study. Journal of Allergy and Clinical Immunology. Elsevier Inc; 2016 Dec 23;:1–29.

24. Comans-Bitter WM, de Groot R, van den Beemd R, Neijens HJ, Hop WC, Groeneveld K, et al. Immunophenotyping of blood lymphocytes in childhood. Reference values for lymphocyte subpopulations. Journal of Pediatrics. 1997 Mar;130(3):388–93.

25. Ishiguro A, Nakahata T, Matsubara K, Hayashi Y, Kato T, Suzuki Y, et al. Age-related changes in thrombopoietin in children: reference interval for serum thrombopoietin levels. British Journal of Haematology. 1999 Sep;106(4):884–8.

26. Despotovic JM, Grimes AB. Pediatric ITP: is it different from adult ITP? Hematology Am Soc Hematol Educ Program. 2018 Nov 30;2018(1):405–11.

27. Kuhne T, Berchtold W, Michaels LA, Wu R, Donato H, Espina B, et al. Newly diagnosed immune thrombocytopenia in children and adults: a comparative prospective observational registry of the Intercontinental Cooperative Immune Thrombocytopenia Study Group. Haematologica. Haematologica; 2011 Dec 6;96(12):1831–7.

28. Walker RW, W. WALKER Idiopathic thrombocytopenia, initial illness and long term follow up. Arch Dis Child. 1984 Apr;59(4):316–22.

29. Kubota M, Adachi S, Usami I, Okada M, Kitoh T, Shiota M, et al. Characterization of chronic idiopathic thrombocytopenic purpura in Japanese children: a retrospective multi-center study. International Journal of Hematology. Springer Japan; 2010 Mar;91(2):252–7.

